# Bridging Neuroimaging and Neuropathology: A Comprehensive Workflow for Targeted Sampling of White Matter Lesions

**DOI:** 10.1101/2025.06.08.25329217

**Authors:** Nadim Farhat, Jinghang Li, Jacob Berardinelli, Mark Stauffer, Andrea Sajewski, Salem Alkhateeb, Noah Schweitzer, Jin Hecheng, Milos Ikonomovic, Jr-Jiun Liou, Howard J. Aizenstein, Joseph Mettenburg, Tales Santini, Minjie Wu, Julia Kofler, Tamer S. Ibrahim

## Abstract

**Background and Purpose:** White matter lesions are common imaging biomarkers associated with aging and neurodegenerative diseases, yet their underlying pathology remains unclear due to limitations in imaging-based characterization. We aim to develop and validate a comprehensive workflow enabling precise MRI-guided histological sampling of white matter lesions to bridge neuroimaging and neuropathology.

**Methods:** We establish a workflow integrating agarose-saccharose brain embedding, ultra-high field 7T MRI acquisition, reusable 3D-printed cutting guides, and semi-automated MRI-blockface alignment. Postmortem brains are stabilized in the embedding medium and scanned using optimized MRI protocols. Coronal sectioning is guided by standardized 3D-printed cutting guides, and knife traces are digitally matched to MRI planes. White matter lesions are segmented on MRI and aligned for histopathological sampling. This approach is validated in over 100 postmortem human brains.

**Results:** The workflow enables reproducible brain sectioning, minimizes imaging artifacts, and achieves precise spatial alignment between MRI and histology. Consistent, high-resolution MRI data facilitated accurate lesion detection and sampling. The use of standardized cutting guides and alignment protocols reduce variability and improve efficiency.

**Conclusions:** Our cost-effective, scalable workflow reliably links neuroimaging findings with histological analysis, enhancing the understanding of white matter lesion pathology. This framework holds significant potential for advancing translational research in aging and neurodegenerative diseases.

## Introduction

White matter lesions are among the most common aging and neurodegenerative disease biomarkers on brain magnetic resonance imaging (MRI) scans. These lesions appear as hyperintense regions on T2-weighted MRI and hypointense on T1-weighted MRI. Broadly, these lesions can be explained by demyelination, axonal loss, and gliosis [1–5]. Clinically, white matter lesions are associated with aging, vascular risk factors (e.g., hypertension, diabetes), and neurodegenerative disorders [3, 6–11]. An increased lesion burden is strongly linked to adverse clinical outcomes, including stroke, cognitive decline, dementia, and mortality [12, 13]. Despite their clinical significance, they remain poorly characterized at the histological level, limiting their utility for disease understanding and treatment development.

Current studies rely on ultra-high field (UHF) postmortem MRI-guided histopathology to bridge the gap between imaging findings and microscopic pathologies [14–25]. However, postmortem MRI at UHF presents unique challenges. Conventional radiofrequency (RF) coils, optimized for in-vivo imaging, produce inhomogeneous RF excitation for ex-vivo specimens, often necessitating restricted fields of view or brain slab dissection to avoid signal dropout [26, 27]. Additionally, standardized protocols for brain embedding, sectioning, and MRI-histology alignment remain underdeveloped. While customized ex-vivo imaging solutions exist, they typically require brain-specific cutting guides that must be individually molded to each specimen’s morphology [28, 29].

To address these challenges, we present a comprehensive, cost-effective workflow integrating cost-effective reusable 3D-printed cutting guides, optimized agarose embedding, and semi-automated MRI-blockface alignment for precise histopathological sampling of white matter lesions. Our method ensures reproducible brain sectioning, minimizes MRI artifacts, and facilitates high-accuracy registration between postmortem imaging and histology. By standardizing the process from MRI acquisition to tissue sampling, this workflow enhances the translational potential for postmortem studies, directly bridging imaging biomarkers and underlying neuropathology.

## Methods

### Workflow Description

Our methodology follows a systematic, multi-step procedure designed to precisely identify and sample white matter lesions through ultra-high field MRI imaging, section cutting and semi-automated alignment.

Brain Preparation and Embedding: We first prepare the postmortem brain and embed it in agarose gel. This stabilizes the tissue during imaging and protects it from deformation during subsequent sectioning steps. The agarose medium is carefully formulated with a saccharose mixture to optimize imaging quality.

Ultra-High Field MRI Acquisition: The embedded brain undergoes scanning with ultra-high field MRI at 7 Tesla, which enables imaging of detailed brain structures at 370 μm isotropic resolution—a level of detail critical for precise alignment with blockface photographs and white matter lesion detection.

Guided Brain Sectioning: Following MRI acquisition, we section the brain into coronal slabs using our custom 3D-printed cutting guides. Each slab is approximately 9.8 mm thick, with the cutting guides ensuring consistent and accurate sectioning. Each slab surface is photographed to create blockface reference images.

Image Alignment: We spatially align the coronal MRI images to their corresponding blockface photographs using the distinctive patterns created by the cutting guides as reference points. These alignments are digitally preserved to maintain spatial correspondence between imaging and physical specimens.

White Matter Lesion Sampling: The segmented white matter lesions, visible on both T1-weighted and T2-weighted sequences, guide precise histopathological sampling. Our in-house deep learning model automatically identifies these lesions on the T1-weighted sequence, and the pathologist uses these lesion masks with the saved MRI-blockface alignments to target specific regions on the physical brain slabs for histopathological analysis.

### Cost-effective Cutting Guide

The cutting guide consists of enclosure, cutting guide, and sealing lid. The enclosure protects the brain tissue and the embedding medium from deformation during scanning (red part in Figure 1-A). The shape of the enclosure conforms to the inner shape of the receive coil (Figure 1-B) and coarsely approximates the shape of a hemispheric forebrain. The enclosure’s bounding box dimensions are: 210mm in the anterior-posterior direction; 80mm in the medial-lateral direction; 170mm in the superior-inferior direction. The enclosure features 3 mm thick walls and a 3.5 mm thick base. Initially, we 3D printed the enclosure and all other components using a high-density ABS M30™ FDM filament at 0.01-inch layer resolution on a Fortus 450 3D printer (Stratasys USA). However, due to issues with flexing and splitting between the layer lines, we transitioned to using polycarbonate material for all parts. This change provides increased layer adhesion and impact resistance. With the polycarbonate material, the printing time for the enclosure is approximately 9 hours. The enclosure is specifically designed to fit within our in-house developed MRI RF coil [30, 31].

**Figure 1.**
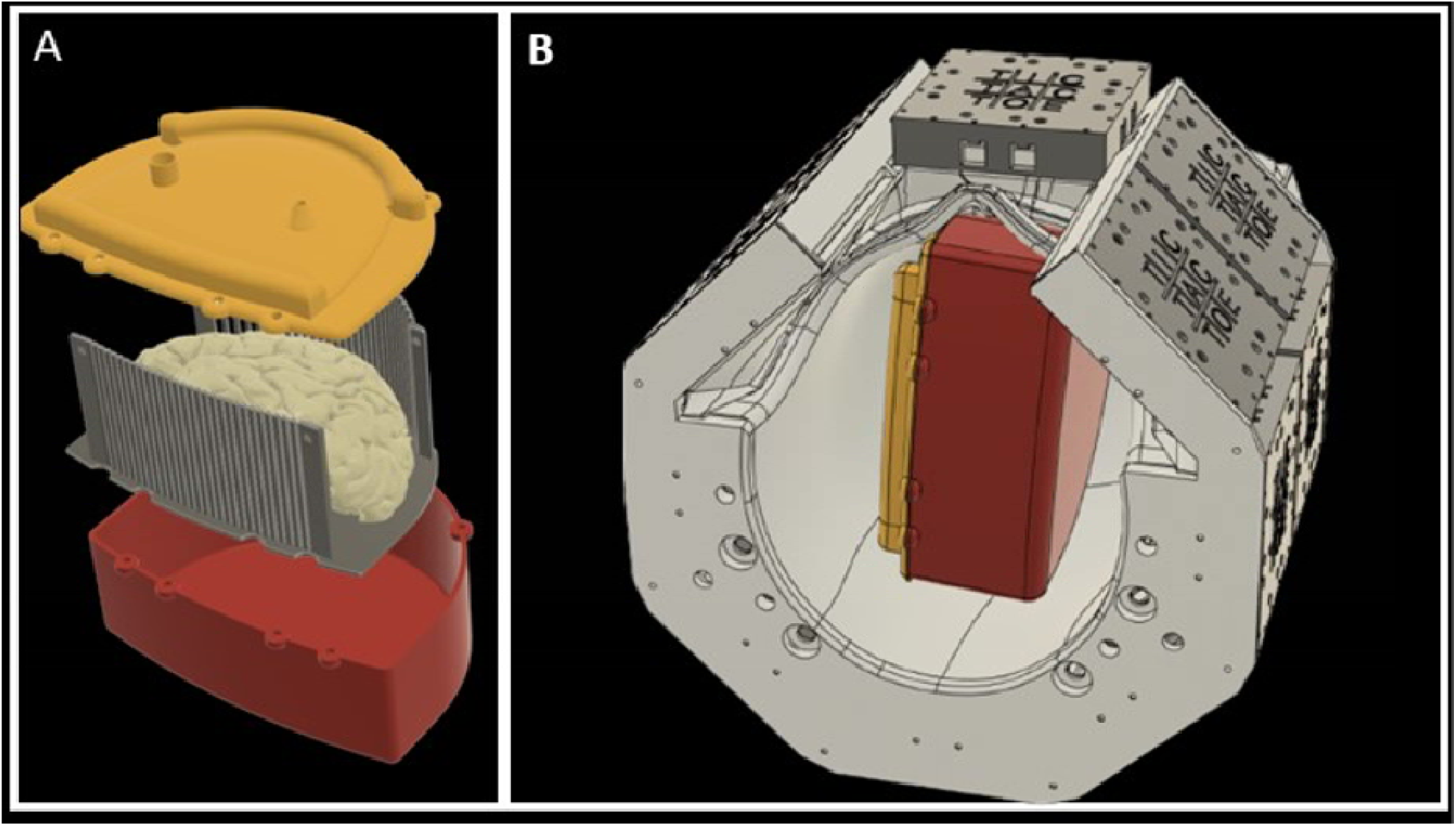
3D Rendering of the 3D printed container with cutting guides. A) The 3D model of the cutting guides (grey), the container enclosure (red), and the sealing lid (yellow). B) 3D rendering shows the fit of the container to the RF coil’s inner dimensions.

The cutting guides (grey part in Figure 1-A) are designed to precisely control brain sectioning by restricting the knife’s movement through the coronal plane. Conforming to the enclosure’s shape, the guide consists of parallel columns with precisely spaced gaps that direct the pathology knife. The structure includes twenty-eight top cylindrical columns arranged in an arc to match the brain’s dorsal curvature, twenty-nine bottom cylindrical columns in a gentle curve to accommodate the temporal cortex, four rectangular corner columns, and a base plate. The cylindrical columns (3 mm diameter) are positioned 0.6mm apart, sized specifically to guide a standard 0.5 mm pathology sectioning knife. These columns span from the prefrontal to occipital lobe, covering the typical extent of a left-hemisphere brain. For fabrication, we 3D-print the cutting guides using high-density polycarbonate FDM filament at 0.005” layer resolution on a Fortus 450 3D printer (Stratasys USA), requiring approximately 22 hours of printing time. In our latest iteration, we transition from polycarbonate to VICTREX AM™ 200 polyaryletherketone (PAEK), significantly extending the operational lifetime of the cutting guide [31].

The sealing lid (yellow part in Figure 1-A) provides a secure seal for the enclosure while incorporating functional features for improved agarose embedding. It features an inlet port for adding additional agarose after the enclosure is closed and an outlet port that enables air suctioning during the filling process. This dual-port design allows for the removal of trappe air bubbles while simultaneously filling the container, improving the quality of the embedding medium. The lid is 3D printed using high-density polycarbonate FDM filament at 0.01“ layer resolution, with a total printing time of approximately 4 hours.

### Brain Embedding

#### Embedding media preparation

To mitigate motion artifacts during scanning and protect the brain from deformation during handling and sectioning, we embed postmortem brains in an agarose-saccharose mixture. This medium not only provides structural support but also improves the signal-to-noise ratio (SNR) and contrast-to-noise ratio (CNR) compared to formalin-fixed brains [32, 33].

For optimal ultra-high-field (UHF) MRI imaging, the embedding media’s dielectric properties must closely match those of the brain. While the relative permittivity of water and agarose is ∼78, the brain’s permittivity is approximately 58 [34]. By adding saccharose to the agarose solution, we reduce the medium’s permittivity from 78 to 68, closely approximating the brain’s dielectric properties at room temperature [15, 35].

#### Media preparation protocol

A solution of 1.5% agarose and 30 % saccharose is prepared by dissolving 0.75g of agarose and 4.5g of saccharose in 50ml water respectively. The agarose-saccharose mixture is then heated to 65-68° Celsius(C) on a hot stir plate over 2-3 hours until fully dissolved, forming a clear solution. After turning off the heat, the solution is continuously stirred at room temperature until cooling to 45-50°C. It is then transferred to a cold room until reaching 37-38°C, at which point it is ready for brain embedding (Figure 2).

**Figure 2.**
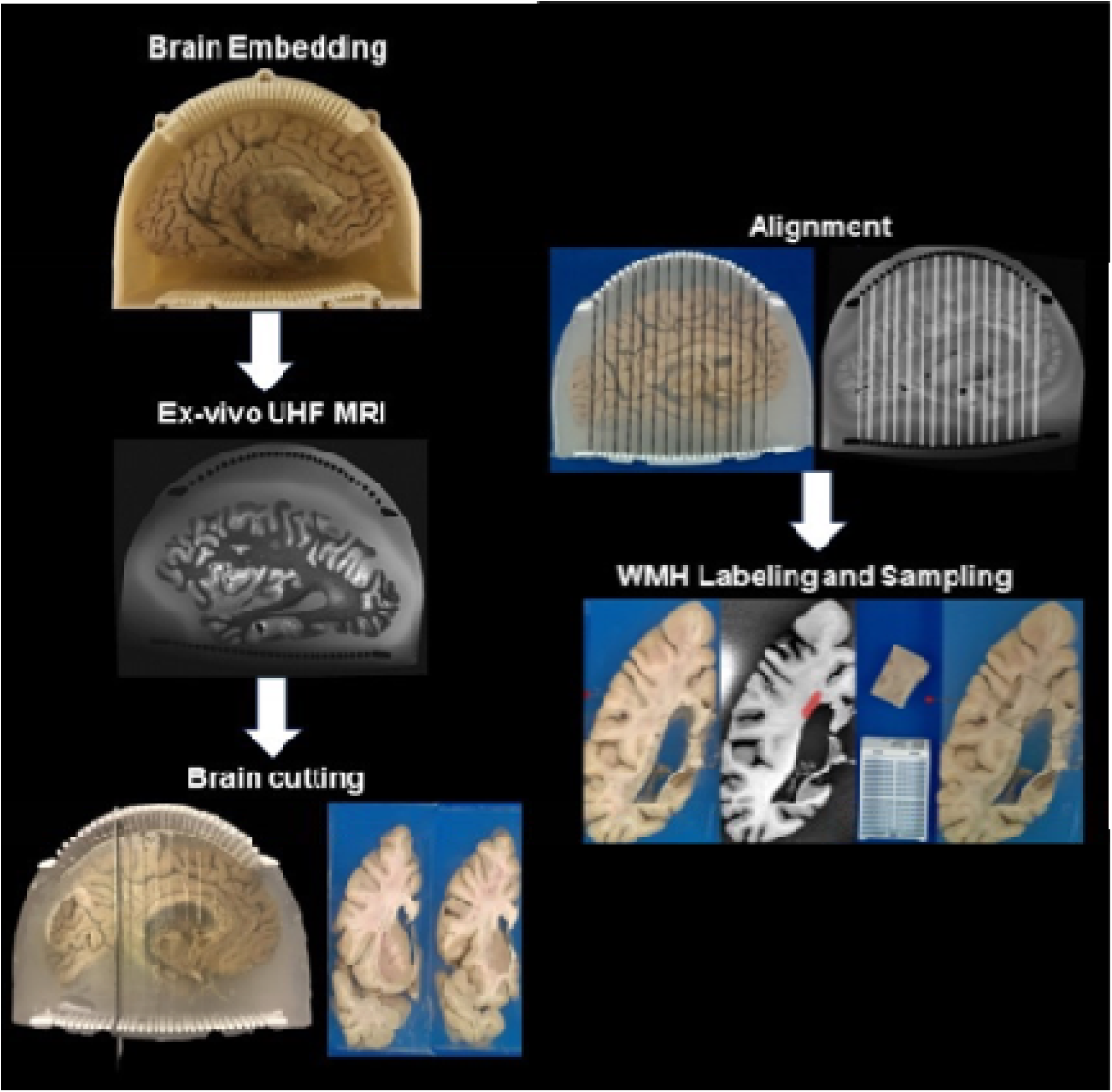
Workflow of the MRI-guided sampling of white matter lesion.

**Figure 3.**
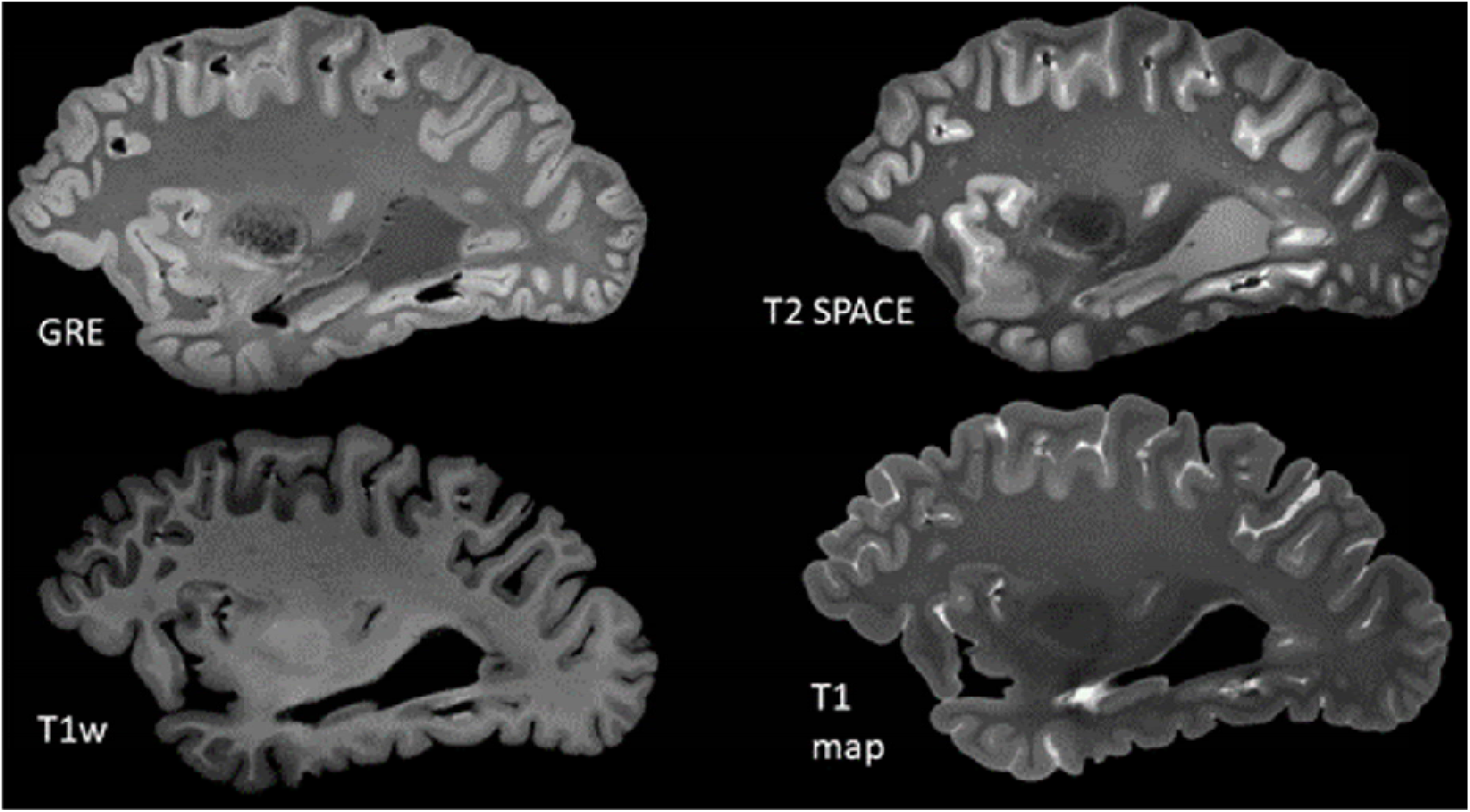
Example of acquired MR contrasts for the postmortem brain MRI.

#### Brain embedding procedure

A cutting guide is placed inside the embedding enclosure, and a 1 cm thick agarose base layer is poured. The left hemisphere of the brain is positioned with the lateral side down and ventricles facing upward. The ventricle is first filled with agarose, followed by careful pouring around the brain while massaging to eliminate air bubbles. The brain is gently pressed into a fully submerged position for 2-3 minutes until the agarose thickened, preventing flotation.

After 2-3 hours, the agarose fully solidifies, forming a stable, transparent encasement. enclosure is then sealed with the lid and is ready for imaging.

#### Postmortem Brain UHF MRI Acquisition

The For scanning, the prepared container is placed inside our custom-designed head coil, as shown in Figure 1-B [36–40]. The brain is imaged using the following MRI pulse sequences: Gradient Echo (GRE): TR = 40 ms, TE1 = 8 ms, TE2 = 16 ms, 256 slices, voxel size = 0.37 × 0.37 × 0.37 mm³, acquisition time = 36 min and 59 s. T2 Sampling Perfection with Application-optimized Contrast using different flip angle Evolution (T2-SPACE): TR = 3400 ms, TE = 368 ms, 208 slices, voxel size = 0.41 × 0.41 × 0.41 mm³, acquisition time = 46 min and 19 s.

Magnetization Prepared 2 Rapid Gradient Echo (MP2RAGE): Two GRE echoes, TR = 6000 ms, TE = 3.57 ms, TI1/TI2 = 556/2200 ms, flip angles = 6°/7°, 240 slices, voxel size = 0.37 × 0.37 × 0.37 mm³, acquisition time = 32 min and 28 s.

### Brain Cutting

Brain sectioning begins by removing the lid of the enclosure (Figure 4A–B). Using the rectangular columns as handles (Figure 4B), the cutting guides are separated from the enclosure by first loosening the embedding medium along the enclosure walls with a knife.

**Figure 4.**
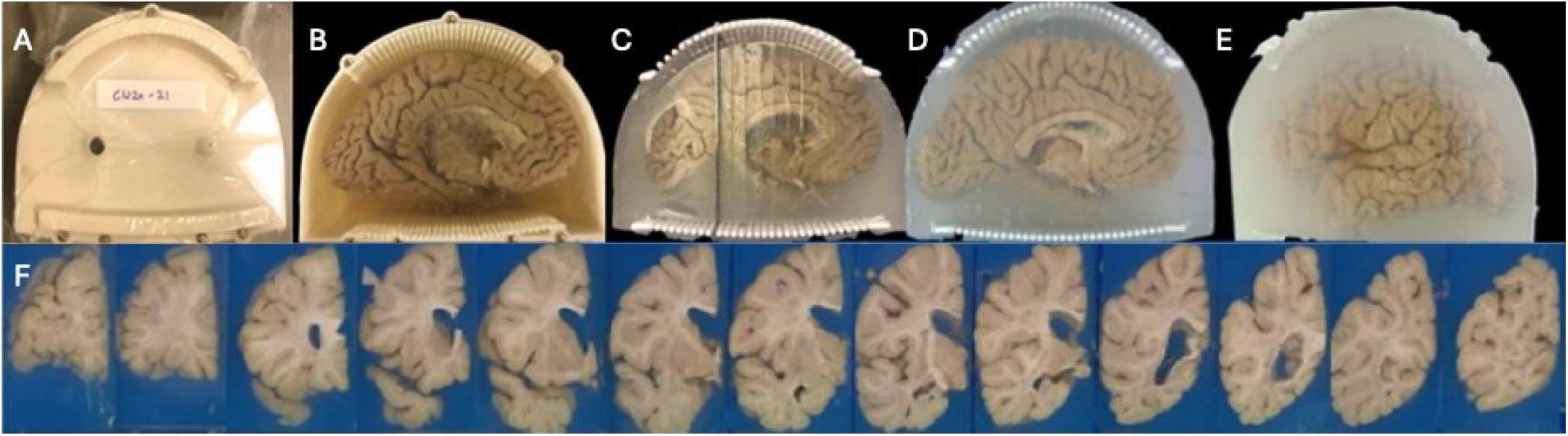
Brain Cutting Process. A) The sealed container with the lid in place. B) After removing the lid, the brain and cutting guide are visible, embedded in agarose. C) The container is removed, and the brain is sectioned using a knife guided by the cutting columns. D) The brain is fully sectioned; both the agarose and brain maintain their structural integrity. E) The cutting guides are removed, leaving the slabs embedded in agarose. F) The slabs are arranged and photographed.

Once detached, the cutting guides along with the embedded brain are removed and placed flat on a working surface.

The neuropathologist starts sectioning by aligning a 12-inch brain knife (CellPath #CAA-1201-01A, 0.5 mm thick) between the top and bottom columns of the guide, corresponding to the planned initial coronal cut (Figure 4C). Subsequent slicing of the left hemisphere brain is performed by inserting the knife between cylindrical columns of the cutting guide—skipping two column pairs per cut—to maintain alignment with the original orientation (Figure 4D). After completing the sectioning, the knife marks on the surface of the embedding medium are photographed. These cut traces are critical for reconstructing the cutting planes and aligning postmortem brain MRI slices during multiplanar reconstruction (MPR) (Figure 4E). Finally, the brain slices are placed in formaldehyde solution for one additional week. During this period, postmortem MRI, and histological data are co-registered to a common space, and regions of interest are identified for downstream sampling.

### Ex-vivo MRI to Blockface Photos Alignment and White Matter Lesion Sampling

Our method aligns ex-vivo MRI with blockface photographs of brain slabs using a fiducial-based approach. In typical fiducial registration—commonly used in neurosurgery—distinct anatomical or artificial markers serve as fixed reference points across imaging modalities to ensure precise alignment [23]. In our protocol, the cylindrical columns of the cutting guides serve this role. These columns are uniquely positioned and easily distinguishable—appearing white in blockface photographs and dark in T2-SPACE MRI images—allowing them to function as reliable fiducial markers (Figure 5). We assume that the knife, when guided between column pairs during slicing, follows a plane normal to the embedding medium. Accordingly, the same plane can be reconstructed virtually in the acquired MR images by identifying the corresponding knife trace between the same column pair.

**Figure 5.**
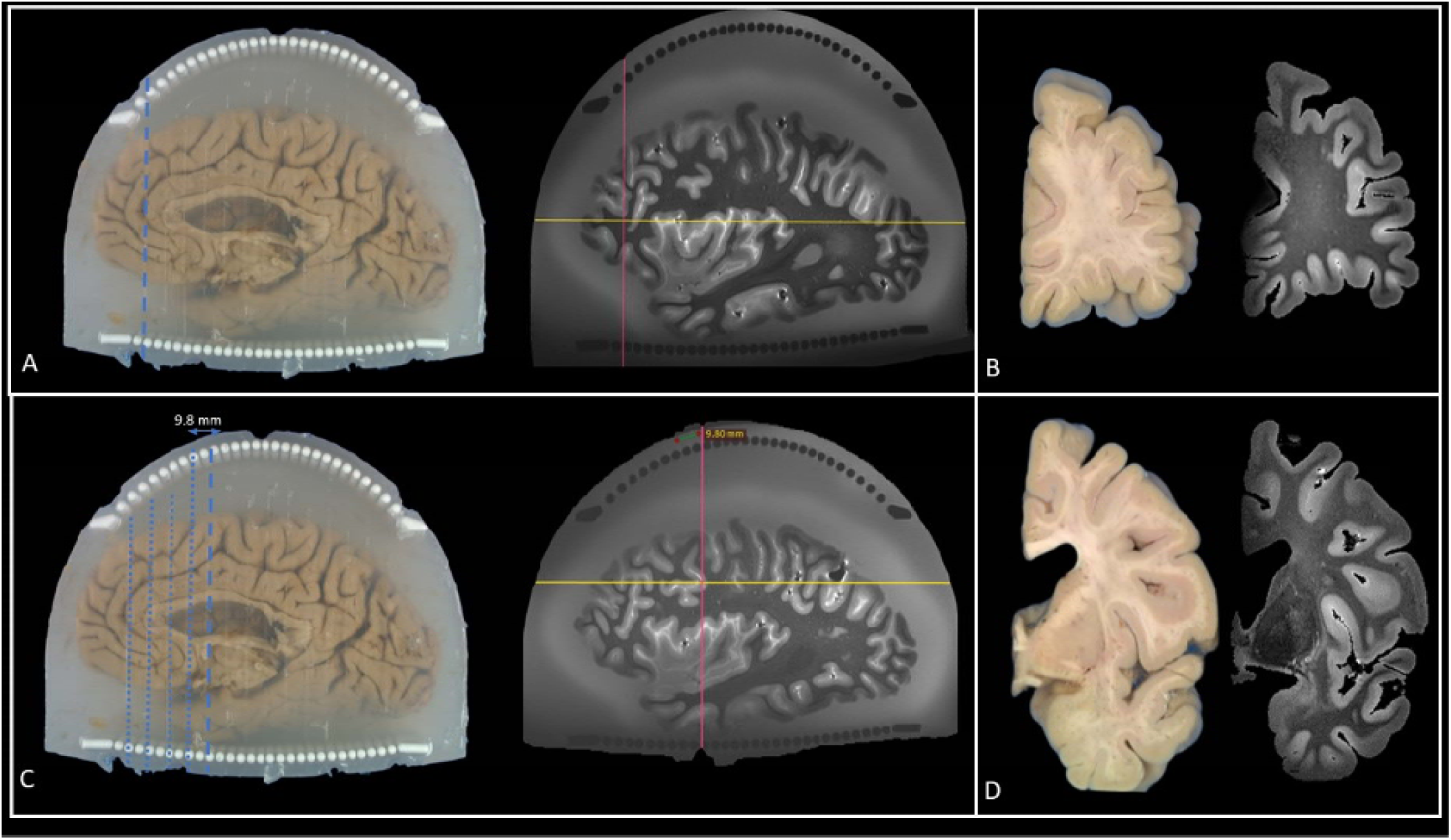
Alignment of MRI virtual cutting planes with knife traces on blockface photographs. A) After sectioning the brain using the cutting guides, the knife leaves visible traces on the surface of the agarose. These traces are manually highlighted w th a dashed blue line. A virtual reconstruction plane (pink line) is aligned to match the orientation of the knife trace and positioned to pass through the corresponding column pair. B) During alignment, each MRI coronal slice is visually matched to the corresponding blockface photograph before proceeding to the next slice. C) As alignment progresses, previously matched knife traces are highlighted in blue, and the current slice position is marked with a blue dashed line. The pink plane denotes the current reconstruction plane in the MRI, aligning with the cutting orientation. D) The visual match between MRI and blockface images are confirmed for each slab before advancing to the next cutting plane.

To perform this alignment, we use the RadiAnt DICOM Viewer (v2022.1.1, 64-bit) with 3D MPR capabilities. The T2-SPACE MRI is loaded into the viewer, and the MPR tool is used to interactively adjust the slicing plane. A corresponding blockface photograph—specifically one showing the first knife trace—is loaded and visually compared. The virtual cutting plane in the MRI is then adjusted to match the knife trace in the photograph, ensuring that the reconstructed MRI slice passes through the same column landmarks. This process is repeated for subsequent slab cuts. Following alignment, white matter lesions are automatically segmented on the MRI and the resulting labels are overlaid on the corresponding blockface images to guide tissue sampling (Figure 6).

**Figure 6.**
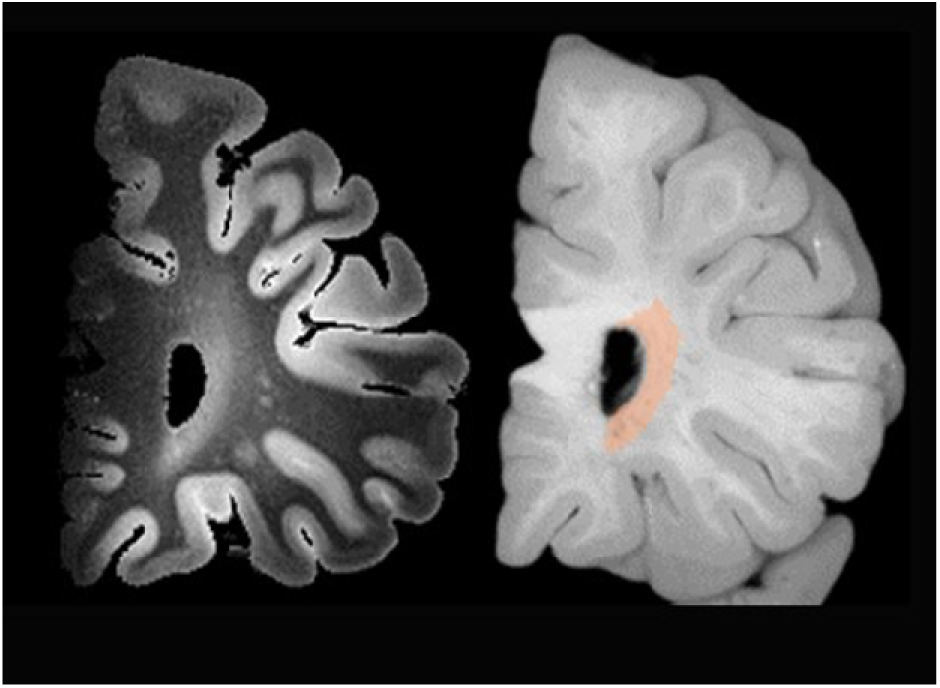
Detection of white matter lesion on T2-SPACE and overlay on a grayscale blockface photo.

## Results

We showcase the alignment results for two postmortem human brains following the workflow illustrated in Figure 2. The brains are first embedded in agarose within the imaging container. Efforts are made to minimize air bubbles in the agarose prior to solidification.

Despite differences in brain size, the embedding medium holds each brain securely in place, and no visible motion artifacts are observed during scanning. Notably, the workflow and pulse sequences remain consistent across both specimens.

Brain 1 is sectioned into 18 coronal slabs—14 using the cutting guides and 4 freehand.

Brain 2 is similarly divided into 18 slabs, with 14 cut using guides and 4 freehand (Figur s 7 and 8). Following sectioning, T2-weighted SPACE images are aligned to their corresponding blockface photographs. Alignment of all 14 guide-assisted slabs typically required ∼30 minutes per brain. In contrast, the 4 non-guided (freehand) slabs required substantially more time due to increased variability (highlighted in yellow in Figures 7 and 8). For both brains, anterior slab surfaces are successfully matched with their corresponding MRI coronal slices across entire left hemisphere. White matter lesions are observed in both brains, spanning periventricular regions from anterior to posterior (solid white arrows, Figures 7 and the the 8).

**Figure 7.**
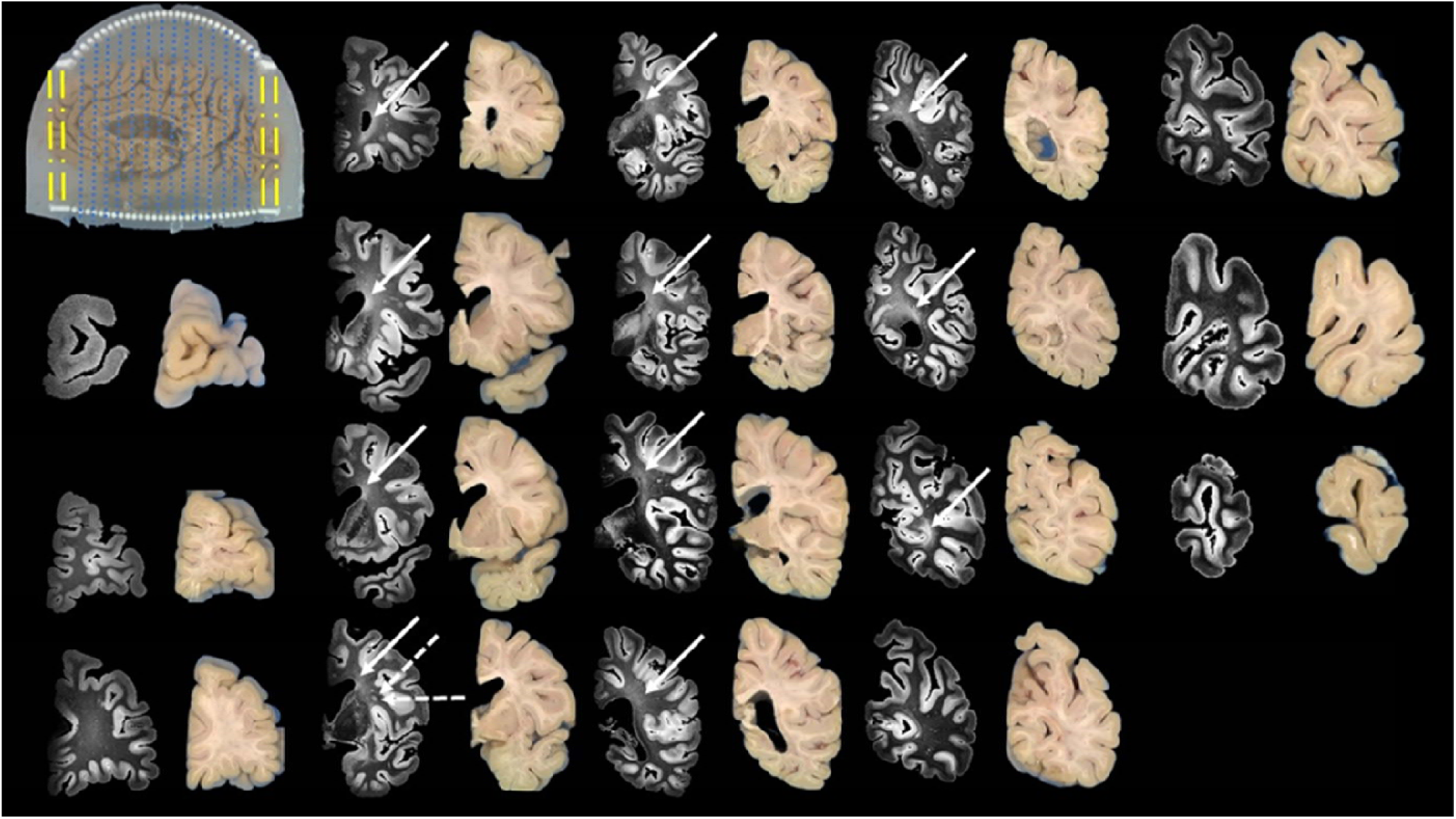
Alignment of T2-SPACE with blockface photos of the brain sample 1. The yellow dashed lines indicate that the first two and last two slices are cut freehand without the cutting guides, and the blue dotted lines indicate that the slices were cut with the cutting guide. The white solid arrows point to the locations of periventricular lesions, and the white dashed arrows point to the location of the deep white matter lesion.

**Figure 8.**
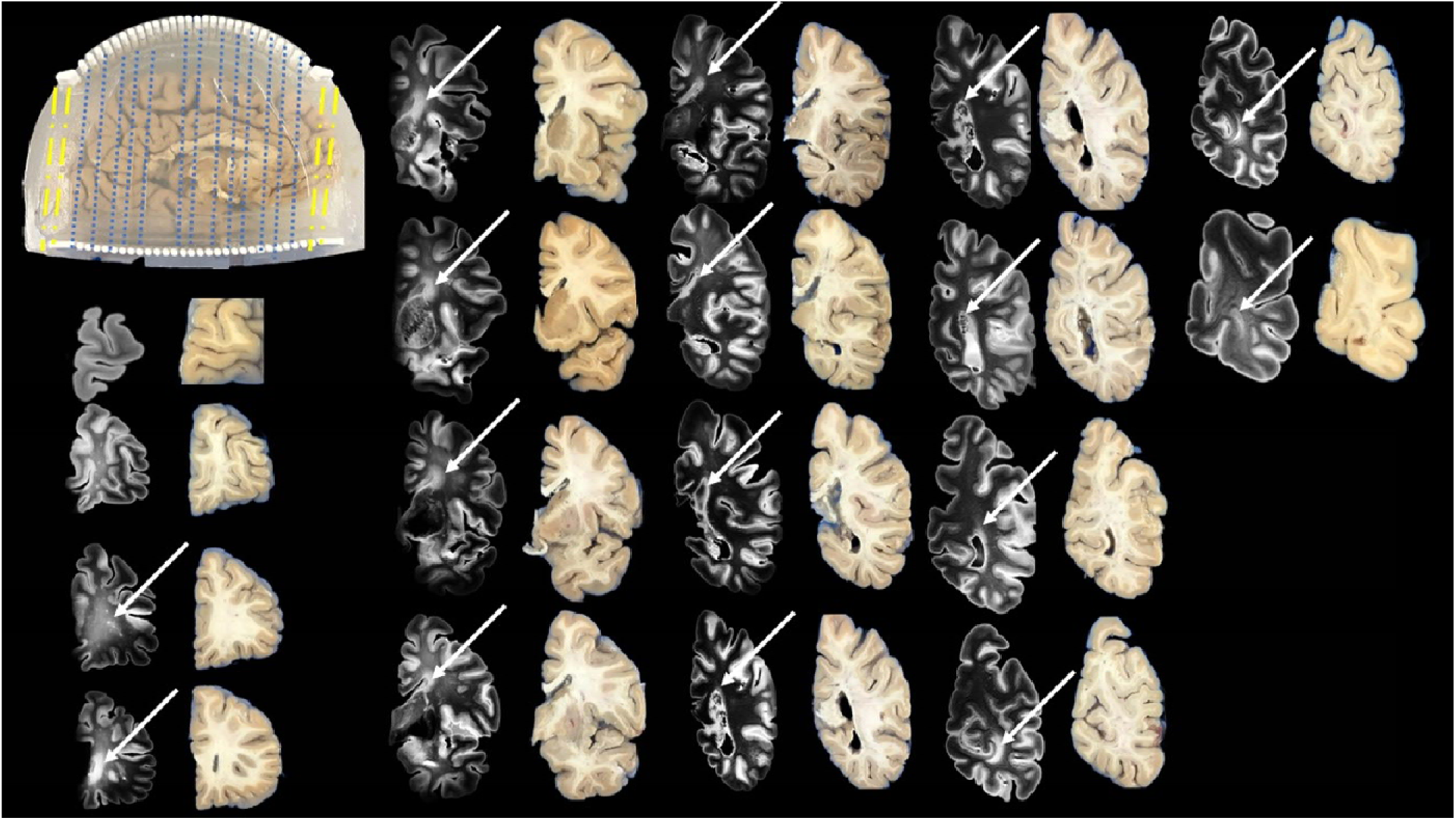
Alignment of T2-SPACE with blockface photos of the brain sample 2. The dashed yellow lines indicate that the first two and last two slices are cut freehand without the cutting guides, and the blue dotted lines indicate that the slices are cut with the cutting guide. The white solid arrows point to the locations of periventricular lesions.

Additionally, Brain 1 exhibits two distinct deep white matter lesion foci (dashed white arrows, Figure 7).

## Discussion

In this work, we introduce a comprehensive, cost-effective workflow designed for precise MRI-guided histological sampling of white matter lesions. By integrating agarose-saccharose embedding, reusable 3D-printed cutting guides, ultra-high-field 7T MRI acquisition, and semi-automated alignment methods, our workflow mitigates inhomogeneity imaging artifacts and achieves precise MRI-histology alignment accuracy. To date, it has been validated over 100 postmortem human brains, demonstrating exceptional reproducibility and scalability.

The agarose-saccharose embedding medium provided exceptional adaptability for various brain sizes by controlling the volume of embedding required based on individual brain volumes. Smaller brains utilized more agarose volume, whereas larger brains required less. Once solidifies, the agarose firmly attaches itself to the brain, cutting guides, and container walls, creating a single, immovable body during MRI scanning, significantly minimizing motion artifacts. This approach enables repeated use of the same container and cutting guides across multiple brains without requiring individualized guides, substantially reducing the cost, especially critical for large scale studies.

Reusable, cost-effective cutting guides plays an integral role in the reproducibility of our workflow. Transitioning from polycarbonate to advanced polyaryletherketone (PAEK) materials notably increases durability, ensuring consistency over repeated usage. During brain sectioning, the agarose embedding secures the cutting guides and brain firmly together, preventing sliding and allowing accurate, stable slicing.

Our workflow’s alignment precision benefits significantly from using cutting guide columns as landmarks for matching MPR planes to the physical knife traces. This efficient alignment process, typically achievable in less than an hour, streamlines the workflow significantly. Furthermore, our container is designed to fit precisely within the inner dimensions of the RF coil, maximizing the space available for postmortem brain loading. Remarkably, none of the brains that we have imaged require exclusion, highlighting the container’s effectiveness in accommodating various brain shapes.

While our agarose-saccharose embedding provides excellent structural support, it occasionally introduces air bubbles, leading to susceptibility artifacts, primarily affecting GRE-based imaging near the sulci and hippocampal regions. Despite technique optimization, these bubbles remain challenging to eliminate fully. Additionally, the reliance on agarose solidification necessitates careful pre-solidification orientation planning, resulting in slight variability in the orientation of the cutting plane across specimens. The cutting guides do not span the entire brain, requiring manual freehand cutting in some areas, which increases alignment complexity and duration. Although container placement within the coil is highly consistent, variability in brain positioning could slightly affect B1+ field distribution.

In conclusion, we develop and validate a systematic, MRI-guided workflow for the precise identification and sampling of white matter lesions in postmortem brains using ultra-high field MR imaging and 3D-printed cutting guides. Our method consists of brain embedding, high-resolution MRI acquisition, guided sectioning, and fiducial-based alignment. The cutting guides significantly improve sectioning consistency. The optimized agarose-saccharose embedding medium minimizes dielectric mismatches and motion artifacts, enhancing imaging quality. Successful applications over 100 postmortem brains demonstrate the reliability and scalability of our approach.

## Data Availability

All data produced in the present study are available upon reasonable request to the authors.

## Acknowledgements and Disclosure

We are deeply grateful to the individuals and families who generously donated their loved ones’ brains for research and science. The 7T imaging acquisitions using the Tic Tac Toe family of RF coils systems were conducted by members of the 7 Tesla Bioengineering Research Program at University of Pittsburgh, under the direction of Dr. Tamer S. Ibrahim. This research was supported by the National Institutes of Health under the grant numbers: R01 AG063525, R01 MH111265, T32 MH119168, T32 HL007560, P01 AG025204, and U19 AG068054. This research was also supported in part by the University of Pittsburgh Center for Research Computing and Data, RRID:SCR_022735, through the resources provided. Specifically, this work used the H2P cluster, which is supported by NSF award number OAC-2117681. The authors declare no commercial or financial conflicts of interest relevant to this study.

